# Preliminary Psychometric Properties of the S-Series Functional Scales (SFS-S, KFS-S, LBFS-S): A Pilot Feasibility Study

**DOI:** 10.1101/2025.10.23.25338612

**Authors:** Ton That Minh Dat

## Abstract

**Background:** Musculoskeletal (MSK) disorders are a leading cause of disability. Existing Patient-Reported Outcome Measures (PROMs) are often long and structurally inconsistent across different joints. We developed the S-Series: a unified system of 6-item, ICF-based, 0-24 point scales.

**Objective:** To report the preliminary feasibility and psychometric properties of the S-Series (SFS-S for Shoulder, KFS-S for Knee, and LBFS-S for Low Back) from a pilot study.

**Methods:** A cross-sectional pilot study was conducted on a convenience sample of 74 participants (22 Knee, 23 Shoulder, 29 Low Back), including healthy controls and symptomatic patients. All participants provided informed consent. Participants completed the relevant S-Series scale and its gold-standard comparator. Feasibility (completion time, ease of understanding), internal consistency (Cronbach’s alpha), convergent validity (Spearman’s rho), and discriminative validity (AUC, sensitivity, specificity) were assessed.

**Results:** Feasibility was excellent. All participants (100%) reported that the S-Series items were “easy to understand” or “very easy to understand.” The average completion time for the entire research packet (which included the 6-item S-Series scale plus its comparator) ranged from 7.1 to 9.7 minutes. Internal consistency was excellent (alpha = 0.92–0.94). Convergent validity was strong (rho = -0.89 to -0.91). Discriminative validity was excellent for KFS-S (AUC = 0.98) and SFS-S (AUC = 0.97), both demonstrating 100% sensitivity. The LBFS-S also showed good discriminative validity (AUC = 0.84) with 85.7% sensitivity and 87.5% specificity.

**Conclusion:** These preliminary pilot data suggest the S-Series functional scales are feasible, reliable, and valid tools for assessing MSK function. The strong psychometric performance supports the rationale for a large-scale validation study, which is currently underway (Hue University IRB Approval No. H2025/627).

## Introduction

Musculoskeletal disorders significantly impact functional independence and quality of life. While established PROMs such as the Shoulder Pain and Disability Index (SPADI) [1], the Oswestry Disability Index (ODI) [2], and the Knee Injury and Osteoarthritis Outcome Score (KOOS) [3] are widely used, they differ in length, structure, and scoring methods. This fragmentation hinders cross-joint comparison and integrated digital assessment.

The S-Series was designed under a unified “4S” framework — Simple, Short, Structured, and Standardized. Each scale contains six core items: three ICF-based (Activity, Participation, Satisfaction) and three symptom-based (Pain, Range of Motion, Strength/Endurance), scored 0–4 each (total 0–24). This structure allows for consistent evaluation across regions and supports both clinical and digital applications.

## Methods

### Study Design and Participants

A cross-sectional pilot study was conducted in July 2025. Seventy-four participants were enrolled: 22 for the knee, 23 for the shoulder, and 29 for the low back. Each group included both healthy and symptomatic individuals.

### Assessment Tools

- **S-Series Scales:** SFS-S (Shoulder), KFS-S (Knee), LBFS-S (Low Back).
- **Comparator PROMs:** SPADI [1], KOOS-12 [3], and ODI [2].
- **Generic Measure:** EQ-5D-5L.

### Feasibility Evaluation

Participants rated the ease of understanding and completion time. Feedback was collected through a brief post-survey questionnaire.

### Statistical Analysis

Following COSMIN guidelines [4]:

- **Internal Consistency:** Cronbach’s alpha.
- **Convergent Validity:** Spearman’s rho between S-Series and its reference PROM.
- **Discriminative Validity:** Receiver Operating Characteristic (ROC) curves, Area Under the Curve (AUC), sensitivity, specificity, and optimal cut-off points.

## Results

### Feasibility

All participants (100%) reported that the S-Series items were “easy to understand” or “very easy to understand.” The average completion time for the entire survey packet (which included the 6-item S-Series scale plus its gold-standard comparator) ranged from 7.1 to 9.7 minutes (Mean ± SD: KFS = 7.1 ± 4.6; LBFS = 8.4 ± 5.3; SFS = 9.7 ± 6.7). This was considered a reasonable time for a research packet.

### Psychometric Properties

The S-Series scales demonstrated strong preliminary psychometric properties, summarized in Table 1. Internal consistency was excellent (alpha > 0.90).

**Table 1.** Preliminary Psychometric Data (n = 74)

| Scale | n | Cronbach's alpha | Comparator (rho) | AUC | Optimal Cut-off | Sensitivity | Specificity |
| --- | --- | --- | --- | --- | --- | --- | --- |
| KFS-S (Knee) | 22 | 0.92 | vs. KOOS-12: -0.89 | 0.98 | $\leq 21$ | 100% | 93.8% |
| SFS-S (Shoulder) | 23 | 0.94 | vs. SPADI: -0.91 | 0.97 | $\leq 22$ | 100% | 83.3% |
| LBFS-S (Low Back) | 29 | 0.92 | vs. ODI: -0.87 | 0.84 | $\leq 21$ | 85.7% | 87.5% |

Convergent validity was strong (rho > -0.89). Discriminative validity was excellent for KFS-S and SFS-S (AUC > 0.97). The LBFS-S also demonstrated good discriminative ability (AUC = 0.84). The optimal cut-off point for dysfunction was identified as ≤ 21 for KFS-S and LBFS-S, and ≤ 22 for SFS-S.

## Discussion and Conclusion

This pilot study provides strong preliminary evidence for the S-Series functional scales. The data show they are highly feasible, reliable, and valid. The excellent discriminative validity (AUC > 0.97 and 100% sensitivity for KFS/SFS) confirms their ability to effectively distinguish between healthy individuals and symptomatic patients.

A key finding was the pronounced ceiling effect observed in healthy control groups (e.g., 75% of healthy KFS/SFS participants scored the maximum 24/24). This is not a limitation, but rather confirms the scales’ intended design: they are highly sensitive tools for capturing dysfunction in symptomatic populations.

The analysis also showed that while the LBFS-S maintained good discriminative ability (AUC = 0.84), its performance was lower than that of the KFS-S and SFS-S. This may be attributed to a more heterogeneous patient group in the low back cohort, including milder cases, which is also reflected in the slightly lower, though still clinically strong, sensitivity (85.7%) and specificity (87.5%) at the ≤ 21 cut-off.

Despite the limitation of a small, convenience sample, these promising results provide a robust rationale for the large-scale, multi-center validation study that is currently underway (approved by the Hue University of Medicine and Pharmacy IRB, No. H2025/627).

## Data Availability

The data that support the findings of this pilot study are available from the corresponding author, T.T.M.D., upon reasonable request.

## Notes

### Competing Interest Statement

T.T.M.D. is the developer of the S-Series functional scales. There are no other competing interests to declare.

### Funding Statement

This study did not receive any funding

### Author Declarations

Ethics committee/IRB of Hue University of Medicine and Pharmacy Institutional Review Board (IRB) gave ethical approval for this work (Approval No. H2025/627).

## References

1. Roach KE, et al. Development of a Shoulder Pain and Disability Index. Arthritis Care Res. 1991;4(4):143–149.

2. Fairbank JC, Pynsent PB. The Oswestry Disability Index. Spine (Phila Pa 1976). 2000;25(22):2940–2952.

3. Collins NJ, et al. KOOS-12: A Shorter Version of the Knee Injury and Osteoarthritis Outcome Score. J Orthop Sports Phys Ther. 2016;46(12):1059–1068.

4. Mokkink LB, et al. The COSMIN Checklist for Assessing Methodological Quality of Studies on Measurement Properties. Qual Life Res. 2010;19(4):539–549.

5. World Health Organization. International Classification of Functioning, Disability and Health (ICF). :Geneva: :WHO; 2001.

